# Demarcation line determination for diagnosis of gastric cancer disease range using unsupervised machine learning in magnifying narrow-band imaging

**DOI:** 10.1101/2020.11.03.20189472

**Authors:** Shunsuke Okumura, Misa Goudo, Satoru Hiwa, Takeshi Yasuda, Hiroaki Kitae, Yuriko Yasuda, Akira Tomie, Tatsushi Omatsu, Hiroshi Ichikawa, Nobuaki Yagi, Tomoyuki Hiroyasu

## Abstract

**Objectives:** It is important to determine an accurate demarcation line (DL) between the cancerous lesions and background mucosa in magnifying narrow-band imaging (M-NBI)-based diagnosis. However, it is difficult for novice endoscopists. Our aim was to automatically determine the accurate DL using a machine learning method.

**Methods:** We used an unsupervised machine learning approach to determine the DLs because it can reduce the burden of training machine learning models and labeling large datasets. Our method consists of the following four steps: 1) An M-NBI image is segmented into superpixels (a group of neighboring pixels) using simple linear iterative clustering. 2) The image features are extracted for each superpixel. 3) The superpixels are grouped into several clusters using the k-means method. 4) The boundaries of the clusters are extracted as DL candidates. To validate the proposed method, 23 M-NBI images of 11 cases were used for performance evaluation. The evaluation investigated the similarity of the DLs identified by endoscopists and our method, and the Euclidean distance between the two DLs was calculated. For the single case of 11 cases, the histopathological examination was also conducted and was used to evaluate the proposed system.

**Results:** The average Euclidean distances for the 11 cases were10.65, 11.97, 7.82, 8.46, 8.59, 9.72, 12.20, 9.06, 22.86, 8.45, and 25.36. The results indicated that the specific selection of the number of clusters enabled the proposed method to detect DLs that were similar to those of the endoscopists. The DLs identified by our method represented the complex shapes of the DLs, similarly to those identified by experienced doctors. Also, it was confirmed that the proposed system could generate the pathologically valid DLs by increasing the number of clusters.

**Conclusions:** Our proposed system can support the training of inexperienced doctors, as well as enrich the knowledge of experienced doctors in endoscopy.

## 1 INTRODUCTION

Several studies have reported that magnifying narrow-band imaging (M-NBI) is useful for diagnosing the existence of early gastric cancer and determining its horizontal extent [1], [2], [3]. The detected cancer lesions can subsequently be removed endoscopically using endoscopic submucosal dissection (ESD) [4], [5].

However, ESD exhibits certain matters of concern, such as complications and the risk of non-curative resection (remnants of cancer) [6], [7], [8]. Although this is also the case for other endoscopic procedures, accidental perforation sometimes occurs [9], [10], [11]. Furthermore, the incorrect recognition of the demarcation line (DL) is disadvantageous to patients. When endoscopists underestimate the DL, this may lead to non-curative resection. In contrast, if they overestimate the DL, they may perform ESD containing extensive unnecessary lesions, increasing the risk of perforation or a long procedure time. Therefore, it is important to determine an accurate DL between the cancerous lesions and background mucosa. However, this determination is difficult for novice endoscopists, with the required long-term training and experience.

With the recent development of machine learning (ML) technology, computer-aided diagnosis (CAD) systems are playing an increasingly important role in the medical field and are expected to aid endoscopists with accurate and stable disease detection and diagnosis. In the gastroenterology field, CAD systems for the detection of esophagus cancer, gastric cancer, *H. pylori* infection, and the characterization of colorectal lesions have been developed and validated for clinical use [12], [13], [14], [15].

In this study, we developed a new CAD system that automatically determines the DL on a given M-NBI image through an ML method. The main objectives of the proposed system are to provide the DL candidates for diagnosis of the disease range of early gastric cancer to endoscopists, as well as to support the accurate and stable diagnosis. In recent years, research on image diagnosis using supervised ML such as deep learning has attracted significant interest in medicine owing to its higher classification and prediction performance compared to conventional methods [12], [13], [16], [17]. However, disadvantages exist in obtaining high-performance results in these methods. First, it is necessary to prepare an enormous amount of annotated training data (for example, ‘lesion’ or ‘non-lesion’ labels are provided to each datum by endoscopists) to construct the classification model. These annotations need to be performed by highly experienced endoscopists and are therefore impractical. Second, the diagnosis rationale and criteria are unclear because the available deep learning methodologies are mainly black-box models. It is difficult to extract such knowledge from a deep learning model [14].

In this study, we used a data clustering method to overcome these issues, which is a type of unsupervised ML technique. Given a single M-NBI image, the proposed method divides it into several segments, and then identifies the clusters in which segments with relevant features are densely located.

Consequently, the boundary between the gastric cancer lesions and background mucosa, the DL, can be identified. The diagnosis rationale or criteria can be interpreted following the determination of the DL because we specify the potentially discriminating features of gastric cancer before performing the data clustering. Furthermore, the proposed method does not require any annotations or computational efforts. Thus, the approach can be applied to determine the DL for early gastric cancer diagnosis.

## 2 METHODS

### 2.1 Proposed method

A schematic of the proposed method is presented in **Figure 1**. It mainly consists of the following four steps: (1) image segmentation, (2) feature extraction, (3) segment-based clustering, and (4) identification of DL candidates. First, an input M-NBI image is segmented into a certain number of superpixels, each set of pixels with similar color features. Second, the image features, including the colors and the texture features, are calculated for each superpixel. Third, the superpixels are grouped into a predetermined number of clusters using a data clustering method. Finally, the boundaries of the multiple clusters are extracted as the DL candidates and are presented to the users of the proposed system (endoscopists). The majority of data clustering methods divide the dataset into clusters to minimize the intracluster distance or/and maximize the intercluster distance. Therefore, we hypothesized that the DL could be extracted as the boundary of the clusters because the cancer lesions that would have different image features from the non-cancer lesions should form the cluster. Note that our system was designed to identify only the DL ‘candidates’, so it does not require image annotation by endoscopists. The final decision of selecting the DL from the candidates is the responsibility of the user endoscopists. The details of each of the four steps are described in the following sections.

**Figure 1.**
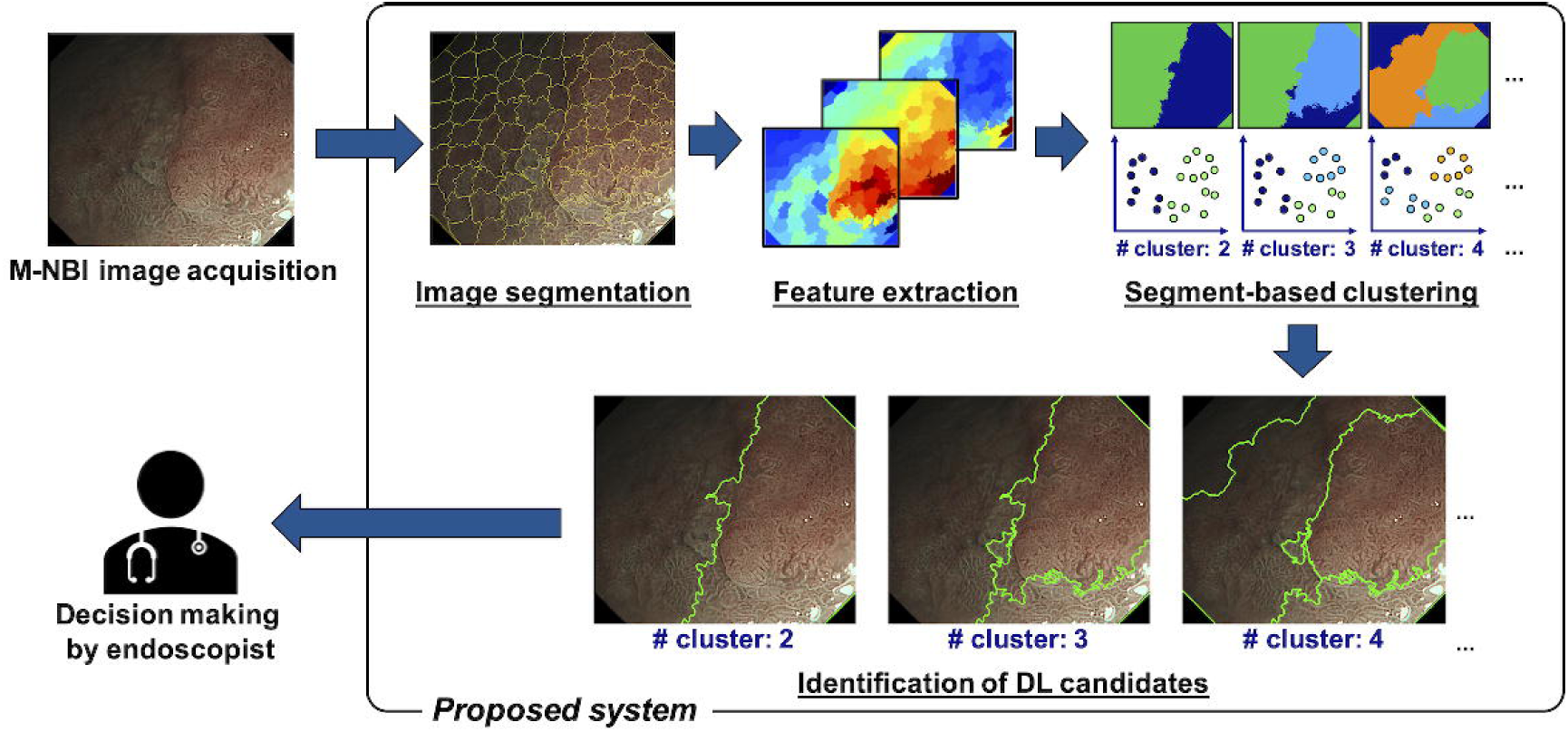
A framework of the proposed method.

#### 2.1.1 Image segmentation

Several superpixel algorithms have been presented that partition an image into multiple segments. In this study, we used simple linear iterative clustering (SLIC) [18], which is one of the most popular methods in image processing. In the SLIC algorithm, a group of pixels that are similar in terms of both their color features and position (coordinates) is formed as a superpixel. The boundary shapes of the superpixels are iteratively refined by means of the well-known *k*-means-based clustering approach. First, the input image is divided into a certain number of grids. The center of each tile (the initial form of a superpixel) is sampled and then moved to the pixel with the lowest color gradient among the neighboring pixels. Thereafter, for each pixel in the entire image, the nearest center pixel in terms of both the color and coordinate spaces is searched, and it is assigned to the member of the identified pixel. This procedure is repeated until certain types of termination criteria are satisfied, so that the superpixels are generated. For further details on SLIC, please refer to the original paper [18]. In this study, we used the *slic* function implemented in scikit-image, which is a Python library.

Prior to applying the SLIC image segmentation, we had to determine the color space to represent the image feature. In this study, we employed the International Commission on Illumination (CIE) L*a*b* color space as the image features for segmentation because it is device independent and also matches with human color perception. It is represented by three-dimensional space consisting of L*, which represents the darkness to lightness, a*, which varies from red to green, and b*, which varies from blue to green [19].

#### 2.1.2 Feature extraction and clustering

After segmenting an input M-NBI image into a set of superpixels, they are further merged into the clusters of the superpixels using the *k*-means method. That is, the local clustering of the pixels to extract the pixel-level image feature is performed by SLIC, following which the global clustering of the superpixels is further performed by the *k*-means method. In the global clustering process, the aim is to identify a cluster of cancer lesions by evaluating each superpixel in terms of both the color and texture features.

The CIE L*a*b* and HSV color spaces were used for representing the color features. The HSV color space is defined by three components: hue, saturation, and value. Hue represents the color type (for example, red, blue, and yellow) ranging from 0 to 360 degrees, saturation represents the color vividness ranging from 0 to 100%, and value represents the color brightness from 0 to 100% [20]. The coordinates in both color spaces can be calculated by nonlinear transformation from the RGB color space.

In addition to the color features, we aimed to extract the texture features of gastric cancer lesions. Yao et al. [21] set their criterion for early gastric carcinoma as (1) the presence of an irregular microvascular pattern or (2) the presence of an irregular microsurface pattern, both of which were accompanied by a clear DL. Therefore, we expected that the texture features of a gastric cancer lesion could be extracted if the irregularities of the microvascular and microsurface patterns were quantified. We used the local entropy, which quantifies the complexity such as the randomness of the image, proposed by Wu et al. [22]. The local entropy score *H*(*S*) of the image block *S* (corresponding to a superpixel in this study) in a given image can be defined by:

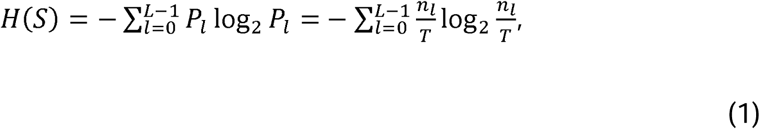

where *l* ∈ {0, 1, …, *L* − 1} denotes the pixel intensity scale (for example, *L* = 256 for an 8-bit grayscale image) and *n*_*l*_, *T*, and *P*_*l*_ denote the total number of pixels within S at pixel intensity *l*, the total number of pixels, and the probability of a pixel having the intensity *l*, respectively. In brief, the entropy used in this case quantifies the variability of the pixel values in the image. That is, we assumed that the irregularities of the microvascular and microsurface patterns could be represented by the probability distribution of the pixel values. Note that we calculated the local entropy score for each color dimension.

In total, we used 12-dimensional feature vectors (that is, the six color dimensions L*, a*, b*, H, S, and V and the local entropy score for each of the six color dimensions) for the global clustering by the *k*-means method. We used the *KMeans* function implemented in scikit-learn.

#### 2.1.3 Identification of DL

Following the global clustering of the superpixels, the boundaries of the clusters are extracted as the DL candidates and are presented on the graphical user interface for the user endoscopists. As mentioned previously, our system is designed to provide the DL candidates and not to determine the exact DL. The final decision on the DL determination is made by the endoscopists by selecting one of the candidates. In this situation, it is necessary to provide varieties of candidates for the users. Therefore, global clustering with different numbers of clusters (the control parameter *k* of the *k*-means method) was executed to provide the DL candidates at different segmentation levels of the M-NBI image. Examples of the cluster boundaries identified following the global clustering are illustrated in **Figure 2**.

**Figure 2.**
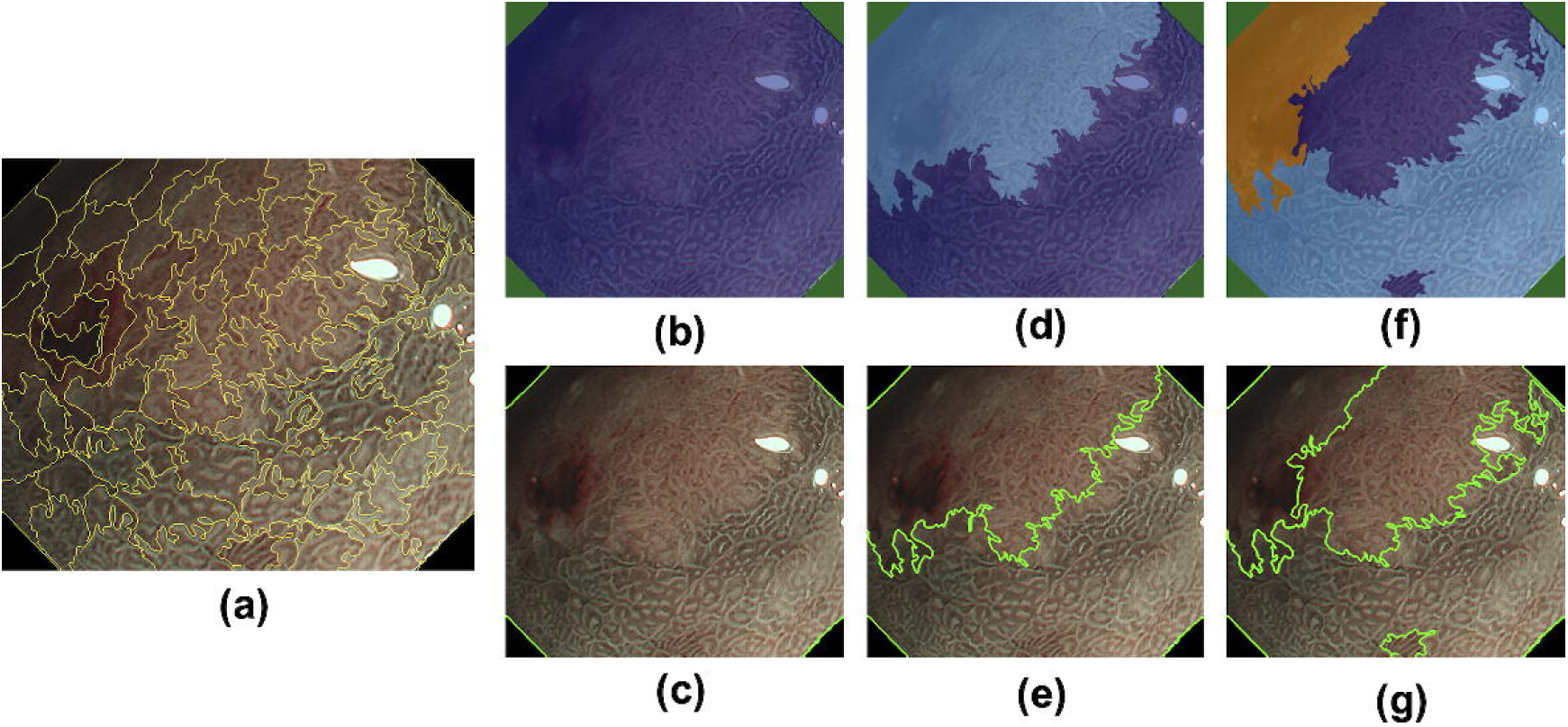
Examples of cluster boundaries identified after global clustering: (a) superpixels identified by local clustering, (b) clusters of superpixels (*k* = 2), where each cluster is painted with a different color, (c) cluster boundaries (*k* = 2), (d) clusters of superpixels (*k* = 3), (e) cluster boundaries (*k* = 3), (f) clusters of superpixels (*k* = 4), and (g) cluster boundaries (*k* = 4).

### 2.2 Experimental setup

To evaluate how accurately our system could provide the DL candidates, we compared them with DLs identified by endoscopists.

#### 2.2.1 Test images

A total of 23 gastric M-NBI images of 11 cases captured at the Asahi University Hospital, Gifu, Japan, from October 2011 to December 2015 were used for the performance evaluation. Those images were anonymized for the sake of personal information is not specified. On the occasion of using still images in this study, all patients provided their written informed consent. Further, the opt-out method was adopted for this retrospective study. The study was approved by the ethics committee of Asahi University Hospital (registration number: 2020-05-01) and was conducted in accordance with the Helsinki Declaration of the World Medical Association and the Ethical Guidelines for Medical and Health Research Involving Human Subjects established by the Ministry of Health, Labour and Welfare, JAPAN.

All examinations were carried out using the EVIS LUCERA or EVIS LUCERA ELITE system and the upper gastrointestinal endoscopes GIF-FQ260Z or GIF-H290Z (Olympus Medical Systems, Co., Ltd., Tokyo, Japan). These endoscopic systems have normal white light observation and NBI modes. All images were acquired in the NBI mode combined with magnifying endoscopy. The clinical characteristics of the patients with early gastric cancer are summarized in **Table 1**. The resolutions of these images were summarized in **Table 2**. Moreover, board-certified gastroenterologists at Asahi University Hospital had drawn the DLs on these images. An example of a DL drawn by an endoscopist is presented in **Figure 3**.

**Table 1.** Clinical features of patients included in test images

| Characteristic | Value |
| --- | --- |
| Male/female, *n* | 6/5 |
| Age, mean, (range), years | 74 (59–92) |
| Tumor size, Average, (range), mm |  |
| Major axis | 14.5 (5-30) |
| Minor axis | 8.8 (4-15) |
| Tumor location, *n* |  |
| Upper | 1 |
| Middle | 3 |
| Lower | 7 |
| Morphologic types, *n* |  |
| Type 0–I | 0 |
| Type 0–IIa | 3 |
| Type 0–IIb | 2 |
| Type 0–IIc | 6 |
| Type1-4 | 0 |
| Depth of tumor, *n* |  |
| T1a (mucosa) | 10 |
| T1b (submucosa) | 1 |
| T2 (muscularis propria) or more | 0 |
| Histological classification, *n* |  |
| Differentiated type | 11 |
| Undifferentiated type | 0 |
| Helicobacter pylori (H.pylori) status |  |
| Current infection | 3 |
| Past infection | 8 |
| Atrophic border |  |
| Closed type | 2 |
| Open type | 9 |

**Table 2.** Resolutions of the test images

| CASE | Image size (pixels) |
| --- | --- |
| | Width ($\mu \pm \sigma$) / Height ($\mu \pm \sigma$) |
| 1 ($n = 4$) | $886.8 \pm 2.2$ / $767.8 \pm 1.7$ |
| 2 ($n = 3$) | $887.0 \pm 1.7$ / $768.0 \pm 1.7$ |
| 3 ($n = 4$) | $1041.0 \pm 0.0$ / $901.8 \pm 0.5$ |
| 4 ($n = 5$) | $887.0 \pm 1.2$ / $767.6 \pm 1.3$ |
| 5 ($n = 1$) | $816.0 \pm 0.0$ / $692.0 \pm 0.0$ |
| 6 ($n = 1$) | $832.0 \pm 0.0$ / $760.0 \pm 0.0$ |
| 7 ($n = 1$) | $799.0 \pm 0.0$ / $691.0 \pm 0.0$ |
| 8 ($n = 1$) | $839.0 \pm 0.0$ / $720.0 \pm 0.0$ |
| 9 ($n = 1$) | $752.0 \pm 0.0$ / $602.0 \pm 0.0$ |
| 10 ($n = 1$) | $800.0 \pm 0.0$ / $650.0 \pm 0.0$ |
| 11 ($n = 1$) | $717.0 \pm 0.0$ / $515.0 \pm 0.0$ |

**Figure 3.**
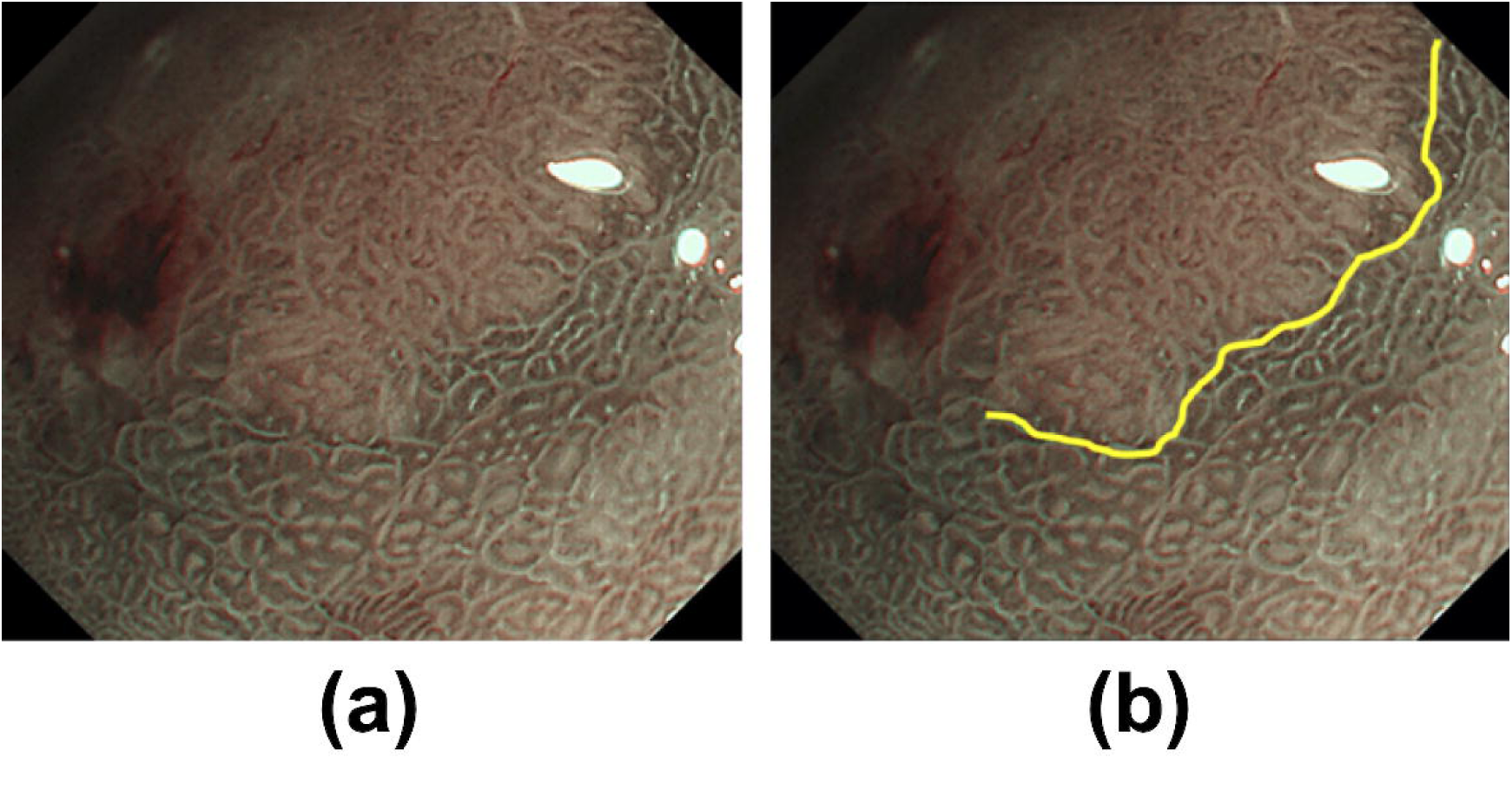
Image with DL drawn by experienced endoscopist: (a) raw M-NBI image and (b) M-NBI image with DL.

Here it should be noted that the pathological evaluation was carried out using the CASE-11 image. The histopathological image of the resected specimen of CASE 11 was also captured and overlapped with the M-NBI image to evaluate whether the proposed method was able to give the boundary between the cancer lesions and background mucosa.

#### 2.2.2 Parameter settings of the proposed method

For the SLIC during the local clustering process, the number of segments (superpixels) of a given image, ‘*compactness*’ parameter, which controls the balance between the color and space proximity, and ‘*sigma*’ parameter, which indicates the width of the Gaussian smoothing kernel for each color dimension, were set to 100, 5, and 5 for L*, a*, and b*, respectively. Moreover, for the global clustering process, the maximum number of iterations of the *k*-means algorithm for a single run was set to 300. The initial cluster centers were determined by the *k-means++* algorithm to accelerate convergence. The execution was carried out with different *k* settings: *k* = 2, 3, 4, 5, and 6. Since the clustering result by k-means depends on the first data point chosen as the centroid for each cluster, it was performed for 10 trials for each k setting, and the best clustering result that indicated the minimum value of the inertia, sum of squared distances of samples to their nearest cluster center, was chosen as the DL candidate.

#### 2.2.3 Performance metrics

We aimed to investigate the similarity between the DLs identified by endoscopists and the proposed method. Therefore, we calculated the Euclidean distance from each pixel of the DL provided by the endoscopist to the nearest pixel of the DL candidates identified by the proposed method as a performance index. All of the image sizes were adjusted to 564×515 to enable a fair comparison in the Euclidean distance calculation.

## 3 RESULTS

**Figure 4** presents the DL candidates provided by the proposed method with different settings of the number of clusters (*k* = 2, 3, 4, 5, and 6) for each of the 23 M-NBI images obtained from 11 cases. The leftmost image includes the DL provided by the endoscopist. The Euclidean distance between the true DL and DL candidates provided by the proposed method is indicated below each image at different *k* values.

**Figure 4.**
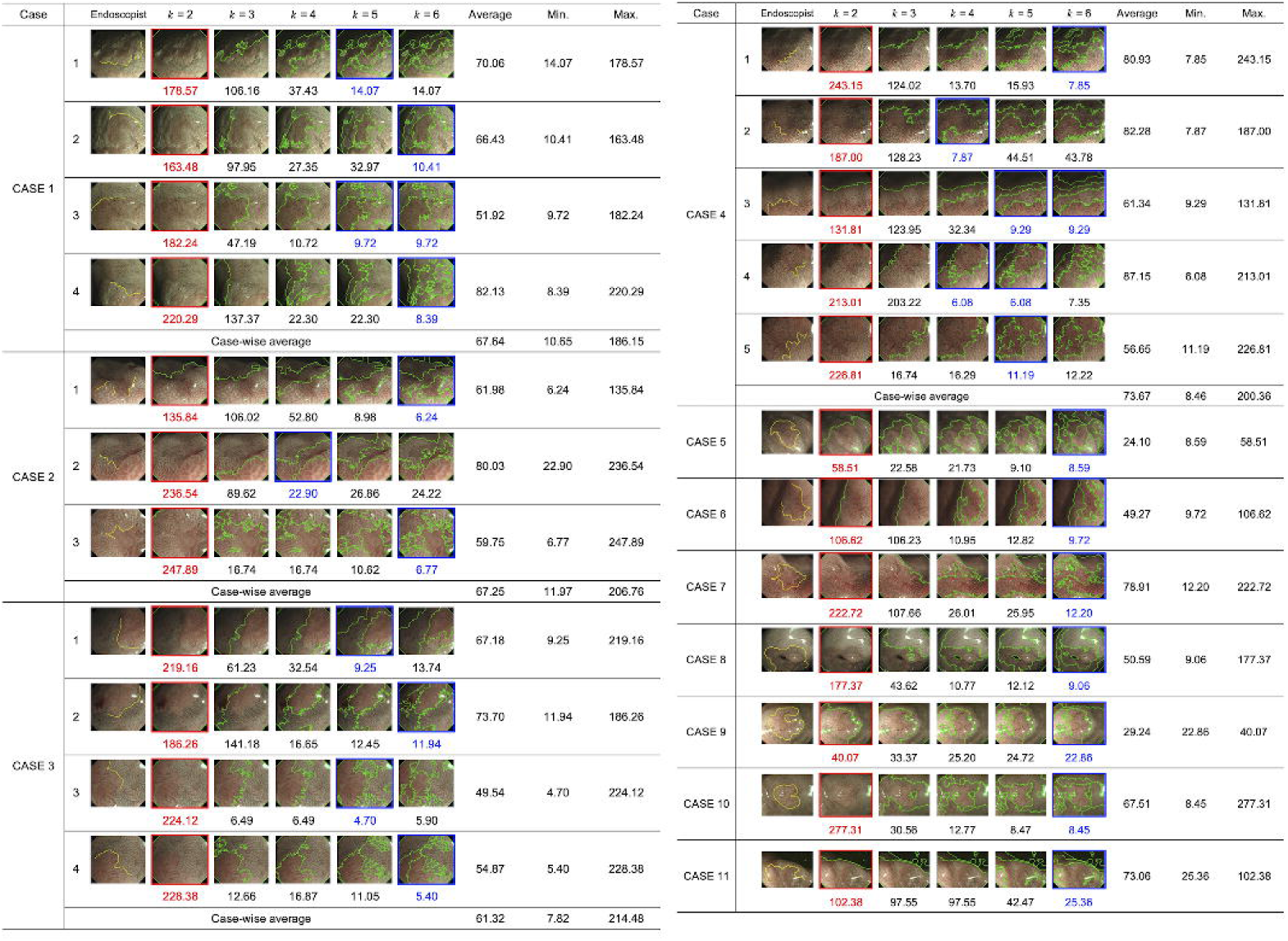
DL candidates provided by the proposed method at different cluster number settings (*k* = 2, 3, 4, 5, and 6) for 23 M-NBI images obtained from 11 cases. The leftmost images present the DLs provided by the endoscopists. The Euclidean distance between the true DL and DL candidates provided by the proposed method is indicated below each image at different *k* values. The average, minimum, and maximum distance was calculated with different *k* settings for each case.

Moreover, the images including the nearest DL candidate to the DL drawn by the endoscopist (that is, where the proposed method provided the best performance) are enclosed by blue lines, whereas the maximum ones are enclosed by red lines. Notably, the average minimum Euclidean distance for CASE 1, CASE 2, CASE 3, and CASE 4 were 10.65, 11.97, 7.82, and 8.46, respectively. Also, the minimum Euclidean distance for the single-sample cases: CASE5, CASE6, CASE7, CASE8, CASE9, CASE10, and CASE11 were 8.59, 9.72, 12.20, 9.06, 22.86, 8.45, and 25.36, respectively.

**Figure 5** indicates the dependence of the Euclidean distance between the DLs provided by the endoscopists and the proposed method on the number of clusters. The dashed black lines correspond to all of the images for all cases. The average distance for all images at each *k* value was calculated and is plotted with a blue point, and these points are connected with a dashed blue line. The average values for *k* = 2, 3, 4, 5, and 6 were 190.13, 87.12, 20.93, 15.68, and 11.99, respectively.

**Figure 5.**
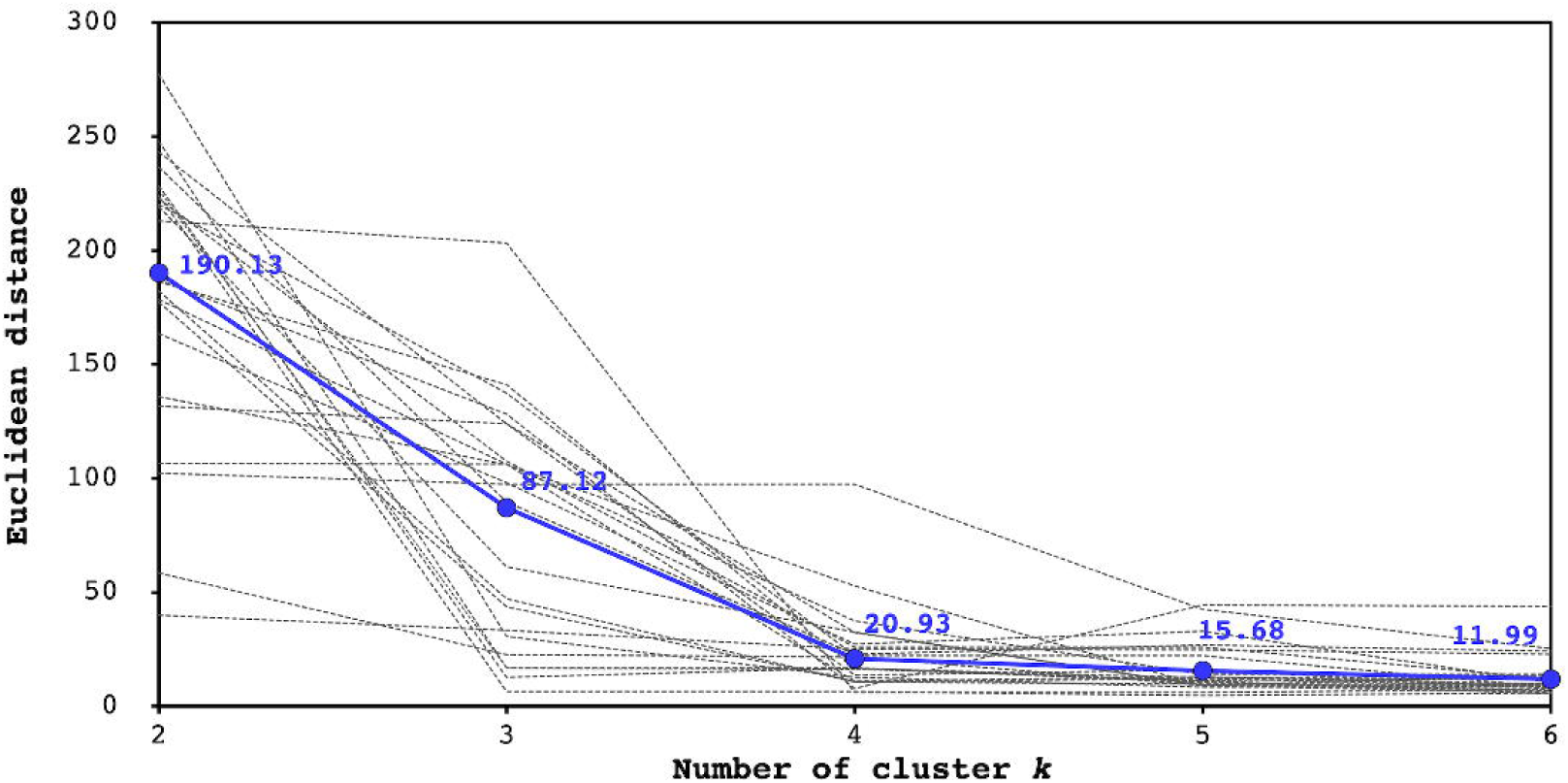
Dependence of Euclidean distance between DLs provided by endoscopists and proposed method on the number of clusters. Each black dashed line indicates the Euclidean distance at each *k* value for each case. The average distance for all the images at each *k* value was calculated and is plotted with a blue point, and the points are connected with a dashed blue line.

To pathologically evaluate the proposed system, the M-NBI image of CASE 11 was overlapped with the histopathological image of the resected sections and the stereomicroscopic image with a similar field of view (FOV) with the M-NBI image, as indicated in **Figure 6. Figure 6(a)** showed the dependence of Euclidean distance between DLs provided by endoscopists and the proposed method on the number of clusters, *k*. **Figure 6(b)** indicated the stereomicroscopic image, overlapped with the hematoxylin-and-eosin (H&E) stained tissue sections #1 and #2 of CASE 11. The cut surfaces #1 and #2 correspond to the cutting lines #1 and #2 shown in **Figures 6(c)** and **(d)**. For **Figure 6(c)**, the cancer lesions on cutting lines #1 and #2 were colored red. The yellow dashed lines were the DLs given by the endoscopists, and the green lines were given by the proposed system. The DLs given by the proposed system in **Figure 6(d)** were the results at *k* = 2 to 20. Also, each cluster was colored differently in **Figure 6(d)**. Furthermore, for *k* = 20 indicated minimum values of the Euclidean distance (**Figure 6(a)**), the clustering results around the cutting lines #1 and #2 were clipped out to observe the shape of clusters and their correspondence to the cancer lesions as shown in **Figure 6(e)**.

**Figure 6.**
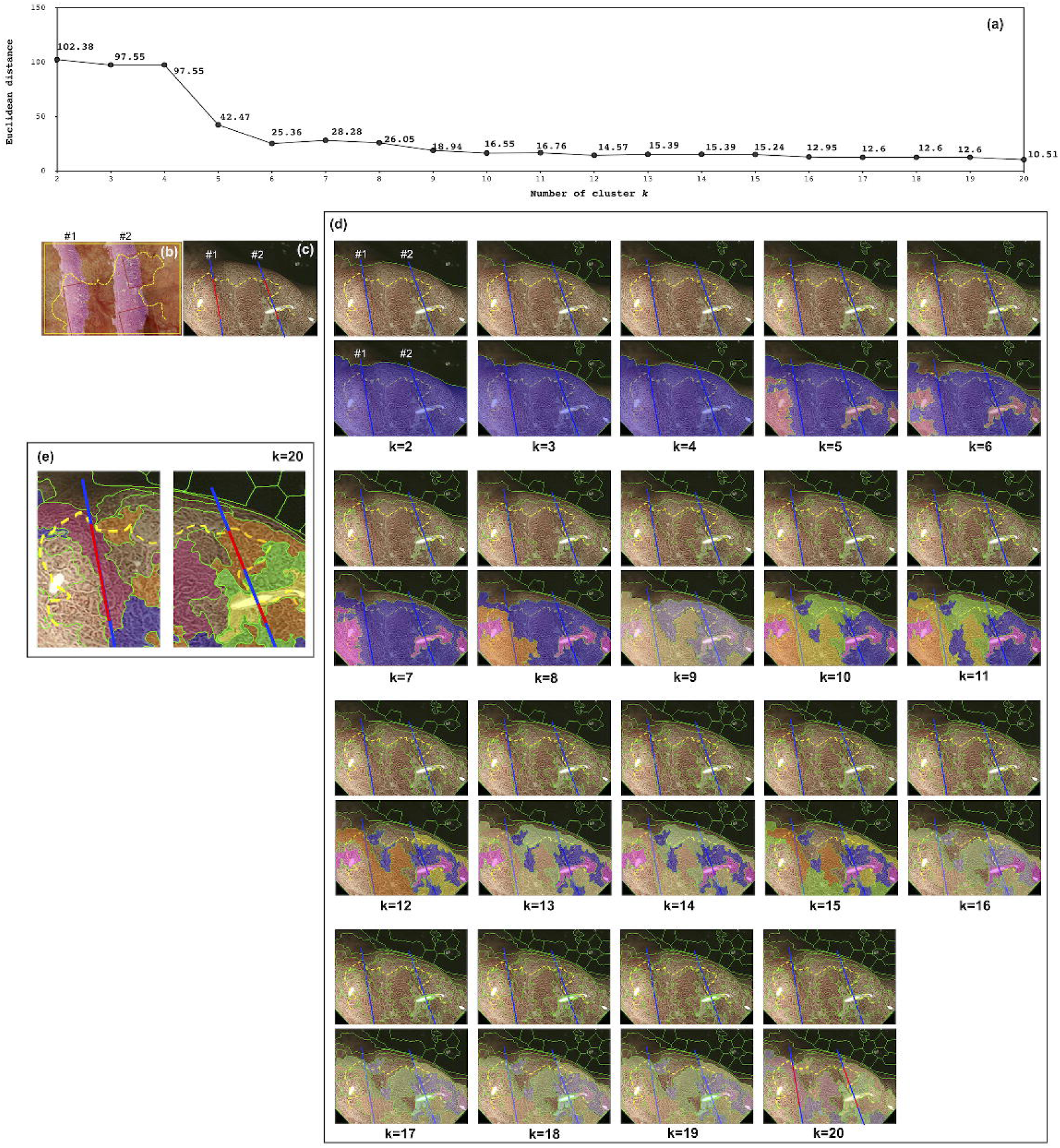
Pathological evaluation of the proposed system for CASE 11. (a) Dependence of Euclidean distance between DLs provided by endoscopists and proposed method on the number of clusters, *k*. (b) shows a stereomicroscopic image of CASE 11 overlapped with the H&E-stained tissue sections #1 and #2 of CASE 11. The cancer lesion is indicated in the red box. The yellow dashed line is the DL provided by the endoscopist. (c) the cancer lesions on cutting lines #1 and #2 were colored red, otherwise colored blue. (d) In addition to (c), the DLs provided by the proposed system for different *k* choices (*k* = 2, 3, …, 20) are indicated by the green lines in the upper figures. For each k, each of the clusters around the cutting lines is shown in a different color from the other clusters in the lower figures. (e) For *k* = 20 in (d), the clustering results around the cutting lines #1 and #2 are clipped out to observe the shape of clusters and their correspondence to the cancer lesions.

## 4 DISCUSSION

### 4.1 Comparison between the endoscopists and the proposed system

It can be observed from **Figure 4** that the different cluster number settings resulted in varying shapes of the DL candidates, and the distance to the DL provided by the endoscopist varied accordingly. Furthermore, **Figure 5** indicates that the DL candidates provided by the proposed method became more similar to the endoscopist identification with an increase in the number of clusters. In most cases, the changes in the Euclidean distance were remarkable when *k* ranged from 2 to 4 and were saturated at *k* >= 4. The approximation accuracy was improved with an increase in the number of clusters because the complexity of the boundary shape increased. These results suggest that the proposed method could identify DL candidates that sufficiently approximated the endoscopist criteria at *k* = 4.

We observed several typical cases of the identified DL candidates to confirm the above assumption. **Figure 7(a)** presents the DL candidates identified by the proposed method for CASE 4. At *k* = 2, the cluster boundary was drawn between the field of view (FOV) and the black corner. At *k* = 3, although the boundary was added in the FOV, it simply divided the FOV into two regions where the brightness of the clusters was differentiated. At *k* = 4, the boundary was finally drawn between the clusters with different mucosal patterns. It can be observed that the DL candidate at *k* >= 4 overlapped the DL provided by the endoscopist. These results visually confirm the assumption that our proposed method could sufficiently approximate the endoscopist criteria at *k* = 4.

**Figure 7.**
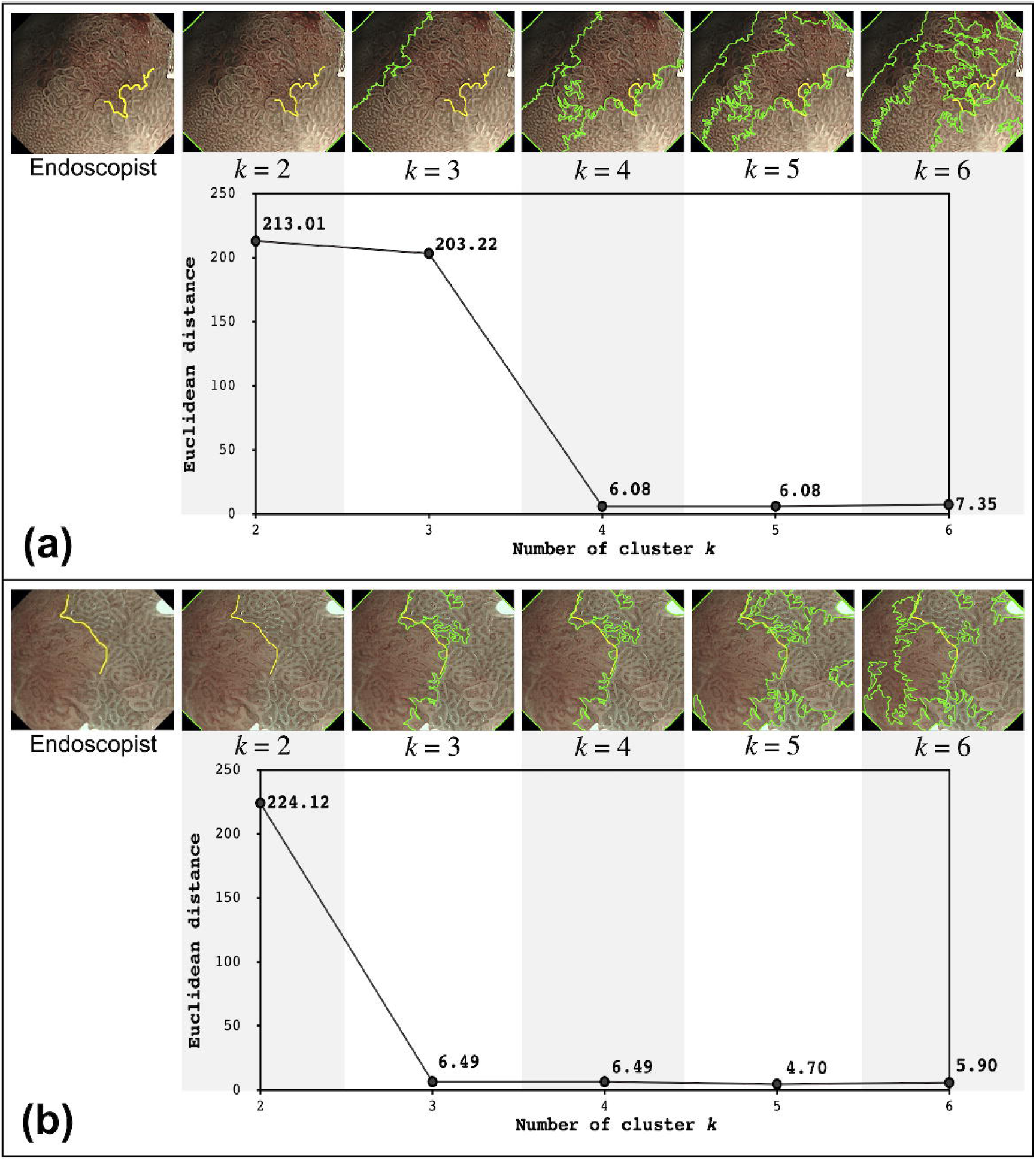
DL candidates identified by the proposed method for (a) CASE 4 and (b) CASE 3. The upper images present the DL candidates at each k value for each subfigure. The yellow and green lines indicate the DLs provided by the endoscopists and the proposed system, respectively. The lower plot shows the dependence of the Euclidean distance between the DLs provided by the endoscopists and the proposed method on different *k* values.

However, as demonstrated by CASE 3 in **Figure 7(b)**, when no shaded regions were observed in the M-NBI image, the proposed method approximated the DL drawn by the endoscopist sufficiently at *k* = 3. No further improvements were observed at *k* > 3, as confirmed both visually and quantitatively in **Figure 7(b)**.

Therefore, it is confirmed that our proposed method could identify the DL similar to that provided by the endoscopist at approximately the number of clusters *k* = 4. Obviously, as this process depends on the varieties of M-NBI images, user endoscopists would select the appropriate one by adjusting the *k* value themselves. However, the observation of multiple DL candidates at different *k* values may offer new insights into endoscopic diagnosis to user endoscopists, because our method can indicate DLs in regions where endoscopists cannot provide a clear demarcation. This is one of the most important contributions of our study.

A further contribution of our study is that it provides the boundary ‘line’ itself, instead of indicating the cancer lesions as a ‘region’. For example, Li et al. [23] divided M-NBI images into non-cancerous and early gastric cancer groups using a convolutional neural network, which is one of the most well-known deep learning models. Moreover, Kanesaka et al. [24] detected cancer lesions using the support vector machine. Although they used a supervised ML method, they also aimed to obtain the cancer lesion ‘region’. Providing a ‘line’ rather than a ‘region’ using a CAD system is meaningful in enriching the knowledge of both inexperienced and experienced endoscopists.

In determining the horizontal extent of early gastric mucosal cancer using M-NBI, it is necessary to identify the DL to differentiate the mucosal patterns between cancer lesions and background mucosa. Consequently, the DL exhibits a complex geometric shape, as illustrated in **Figure 8**. CASE 4 in **Figure 8(a)** included depressed lesions, and the endoscopist indicated the DL on the boundary between the depressed lesion and background mucosa. Moreover, **Figure 8(c)** indicates that the DL was drawn by tracing the light blue crest in CASE 3. In both cases, the endoscopists drew the DLs to trace the structural change in the white zone, and the DLs resulted in an intricate shape being formed. Notably, our proposed system could precisely provide these irregular and complex shapes, as illustrated in **Figures 8(b)** and **(d)**.

**Figure 8.**
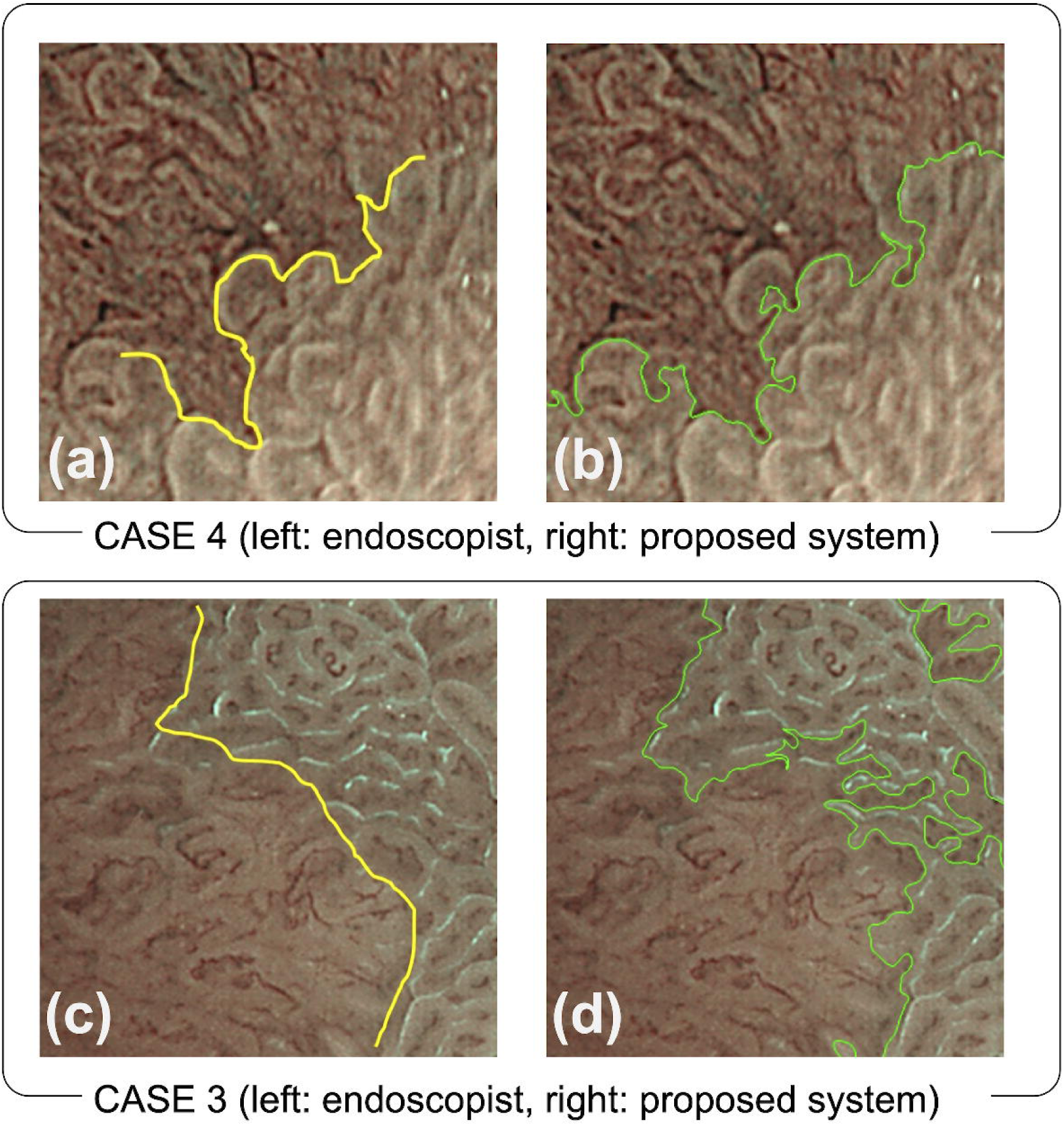
Comparison of DLs provided by endoscopist and proposed method. The top two images (a) and (b) indicate the DLs identified by the endoscopist and proposed system, respectively, for CASE 4, whereas the bottom two images (c) and (d) are for CASE 3.

### 4.2 Pathological evaluation of the proposed system

The proposed system was expected to be able to separate cancerous and noncancerous lesions by cluster boundaries. Therefore, for CASE 11, it was desirable to detect the area corresponding to the red line in **Figure 6(c)** as a single cluster. This assumption was confirmed with the results at k = 20 in **Figures 6(d)** and **(e)**. The red line in cutting line #1, was included in the single cluster as shown in the left figure of **Figure 6(e)**. Also, two cancerous lesions of cutting line #2 indicated by red lines in the right figure of **Figure 6(e)** were part of the orange-colored cluster. It should be noted that this result was not obtained at *k* <= 6 although the dependence of the Euclidean distance has an inflection point at *k* = 6 as shown in **Figure 6(a)**. We concluded that it was due to the existence of the larger shaded regions in CASE 11 than in other cases. The existence of the larger shaded regions in the M-NBI image leads to unnecessary segmentations for them. Even at *k* >=17, the shape of the clusters in the shared regions varied, as shown in **Figure 6(d)**. To overcome this issue, it is necessary to incorporate the predetermination of the regions of interest in the M-NBI images. Nonetheless, it was confirmed that the current system could generate the pathologically valid DL by increasing the number of clusters, *k*.

However, this study exhibits certain limitations. First, the small sample size (*n* = 11) is problematic. Our system needs to be tested with a larger sample size to improve its reliability and robustness. This work focused on the proposal and development of a general framework for an automatic DL determination algorithm based on a data clustering method. Second, each DL may contain an individual bias because a single endoscopist drew it for each image. Although the DLs were assessed by multiple doctors in the present data, the other DL candidates should be provided by various endoscopists to reduce such bias. Thirds, the current study was conducted using only the still M-NBI images. With a recent advance in CAD systems using video images, the future direction should be to investigate the effectiveness of our proposed system using video images. This would be also effective for increasing the sample size. These issues should be addressed for future studies to improve the system reliability and to proceed to clinical trials. However, irrespective of these limitations, the current results demonstrate the effectiveness of our proposed method and will contribute to the development of gastroenterology.

In summary, we developed a new CAD system that can automatically determine the DL on a given M-NBI image through an ML method. In the proposed system, the boundaries of the clusters within M-NBI images are extracted as the DL candidates. The performance evaluation of the system on actual M-NBI images revealed that it can represent the complex shapes of the DLs, similarly to the DLs identified by experienced doctors. The identification of the DL is often quite a difficult task for inexperienced endoscopists. Therefore, our proposed system, which can provide a variety of DL candidates, may support the training of inexperienced doctors, as well as enrich the knowledge of experienced doctors.

## Data Availability

All data relevant to the study are included in the article.

## ACKNOWLEDGEMENT

We would like to thank Editage (www.editage.com) for English language editing.

